# A globally available COVID-19 – Template for clinical imaging studies

**DOI:** 10.1101/2020.04.02.20048793

**Authors:** G.A. Salg, M.K. Ganten, M. Baumhauer, C.P. Heußel, J. Kleesiek

## Abstract

**Background:** The pandemic spread of COVID-19 has caused worldwide implications on societies and economies. Chest computed tomography (CT) has been found to support both, current diagnostic and disease monitoring. A joint approach to collect, analyze and share clinical and imaging information about COVID-19 in the highest quality possible is urgently needed.

**Methods:** An evidence-based reporting template was developed for assessing COVID-19 pneumonia using an FDA-approved medical software. The annotation of qualitative and quantitative findings including radiomics features is performed directly on primary imaging data. For data collection, secondary information from the patient history and clinical data such as symptoms and comorbidities are queried.

**Results:** License-royalty free, cloud-based web platform and on-premise deployments are offered. Hospitals can upload, assess, report and if pseudonymized share their COVID-19 cases. The aggregation of radiomics in correlation with rt-PCR, patient history, clinical and radiological findings, systematically documented in a single database, will lead to optimized diagnosis, risk stratification and response evaluation. A customizable analytics dashboard allows the explorative real-time data analysis of imaging features and clinical information.

**Conclusions:** The COVID-19-Template is based on a systematic, computer-assisted and context-guided approach to collect, analyze and share data. Epidemiological and clinical studies for therapies and vaccine candidates can be implemented in compliance with high data quality, integrity and traceability.

An additional explanation video of the COVID-19-Template video is provided via:http://cloud1.mint-medical.de/downloads/player/index.html?v=Covid19StandardizedAssessmentWeb

**Highlights:**

- Dynamic evidence-based electronic case report form (eCRF) for COVID-19 including documentation of primary imaging data, secondary clinical data and patient history including radiomics features
- Computer-assisted, context-guided reporting approach based on FDA approved medical product software package available free of charge
- Data quality, traceability, integrity in open-access web platform
- Customizable analytics dashboard for explorative real-time data analysis of imaging features and clinical information
- Human and machine-readable data export for clinical trials

## Introduction

*The infection with SARS-CoV-2* has become pandemic since the first cases have been reported from the Hubei province to the WHO on Dec. 31^st^, 2019. This led to a global health crisis with impacts for each individual patient infected with COVID-19, societies and economies.

Dynamic movement patterns, high contagiousness in combination with an estimated substantial asymptomatic and undocumented proportion of infected people^1^ enables a fast spread of the infection and might only be slowed down by drastic disease containment efforts while vaccination and therapy are lacking.^2-4^ The current situation in Italy and other parts of Europe proves that the previous measures for disease control did not suffice and that even countries with a strong health care infrastructure are not adequately staffed with critical care resources. Its current spread in low income countries or those with weak healthcare systems is to be feared.

Besides more aggressive containment, more efforts towards an evidence-based clinical diagnosis, treatment and monitoring including the utilization of new technologies and artificial intelligence are to be strived for.^5^ The global spread of COVID-19 demands a global response across national borders. We perceive an urgent need for systematic data collection, comprising imaging data and analysis, of patients suffering from COVID-19 pneumonia. Therefore, we wish to initiate an open-access platform for data collection using commonly developed criteria for multi-modal diagnostics, staging and monitoring of the disease. This will serve as a mandatory foundation for swiftly conducting future clinical trials aiming at investigating novel diagnostic and therapeutic options.

### Current evidence based on first studies

At present, real-time polymerase chain reaction (rt-PCR) is commonly used for diagnosis of COVID-19. This method, however, requires sophisticated lab technology and material with limited availability. Until now, the results are available earliest the next day. Meanwhile, the patient has to be treated as potentially COVID-19 positive, requiring costly isolation for safety of staff and eventually cohort isolation with a risk to get infected therein. While this method detects the virus, it does not provide quantitative data on the extent or stage of the disease. Fang et al. claimed that the sensitivity of rt-PCR at +3/-3 days after the appearance of external symptoms is only 71%.^6^ The study also found that in 98% of patients at the same stage, abnormalities that are characteristic of viral pneumonia are visible in computed tomography (CT) of the lungs.^6^ The correlation of COVID-19 infection with features in chest CT has been confirmed by other studies.^7^ A sensitivity of 91.9% for a combination of a chest CT and rt-PCR test, compared to 78.2% for a single rt-PCR alone or 86.2% for two subsequent rt-PCR tests was found.^7^ Researchers from China and the Netherlands examined data from 1,014 patients and concluded that CT imaging has high sensitivity for diagnosis of COVID-19 and furthermore offers the capability of screening, comprehensive evaluation, and follow-up.^8^ Based on the chest CT imaging findings it can be concluded that there are correlations between the stage of the disease and the radiological appearance.^9^ The imaging patterns visualized on CT offer the possibility to predict patient progression and the development of potential complications.^9^ Guan et al. demonstrated that 18% of the total patients, who received an initial chest CT (n=877) had no radiological abnormalities (157/877) while only 3% (5/173) of the patients with severe disease course had no radiological abnormalities.^10^ Although there might be a selection bias in many studies, rational usage of chest CT is a powerful tool in clinical monitoring of COVID-19. Moreover, timely underpinning of contagiosity for hospitalized patients via chest CT is crucial to prevent nosocomial infections and exposing medical staff at risk and thus additionally reducing human resources in hospitals. As the severe form of COVID-19 seems to show preference for middle-aged to elderly individuals with comorbidities,^11^ the imaging information could be combined with other patient characteristics to predict the prognosis of patients, so as to identify those requiring intensive medical care.^12,13^ Early stages of COVID-19 pneumonias mainly involve pure GGOs evolving to multiple GGOs with consolidations in lesions and crazy-paving pattern in a progressive stage.^14,15^ This disease monitoring by chest CT imaging has already been used in studies to determine treatment regimens of patients.^11^ We conjecture that the aggregation of radiomics features in correlation with rt-PCR, patient history, clinical and radiological findings, systematically documented in a single database, will lead to optimized application of available resources in terms of diagnosis, risk stratification and response evaluation of COVID-19.

The parameters for systematic data collection were determined based on the current evidence [last update: 03-17-2020] and recommendations from several international medical associations. Findings in the categories patient history and exposure, comorbidities, clinical symptoms, clinical chemistry and radiological findings that might contribute to diagnosis or monitoring of COVID-19 or might be of prognostic value for the course of the disease are included in the developed electronic case report form (eCRF). As comorbidities have been described as a variable with prognostic value, study data for comorbidities such as hypertension^10-13,16-18^, cardiovascular diseases^10-13,16,17,19,20^, chronic obstructive lung disease and other respiratory system diseases^10-13,16,17,19,20^, diabetes^10-13,16-18,20^ and malignant diseases^10-13,17,19,20^ have been pooled (Figure 1, left). Clinical symptoms include fever,^10-13,15-21^ cough,^10- 13,15-21^ expectorations,^10-13,15-19,21^ hemoptysis,^10,11,16,18^ dyspnea,^10-13,15,16,19-21^ headache^10-13,15-21^ and diarrhea^10-13,15-20^ (Figure 1, middle). Radiological findings in chest CT performed on patients with confirmed COVID-19 show radiological abnormalities in the majority of the cases (86.7%, 1122/1294),^10,16,17,19^ typically with bilateral lung involvement,^10,11,17,18^ peripheral distribution^16,17,19,21^ and ground-glass opacification of the lesions ^10,16-19,21^. Depending on the stage of the disease consolidations^16-19,21^, intra-^18,21^ and interlobular^16-18,21^ septal thickening, crazy paving pattern,^16-18^ spider web signs^16,18^ and air bronchogram sign^17,19,21^ were observed (Figure 1, right). In contrast, pleural effusion^16-19,21^ and lymphadenopathy^16-19,21^ were found rarely and do not seem to be typical for COVID-19 pneumonia. Although current evidence for COVID-19 pneumonia relies on data from preliminary clinical studies and thus is most likely subjected to change over time, we illustrated that some clinical symptoms and radiological findings were found more frequently than others, supporting a diagnosis of COVID-19 pneumonia, whereas others seem to be very rare and support a consideration of other differential diagnoses (Figure 1). Nevertheless, or precisely because of it, these parameters were included in the eCRF to enable a solid decision-making.

**Figure 1:**
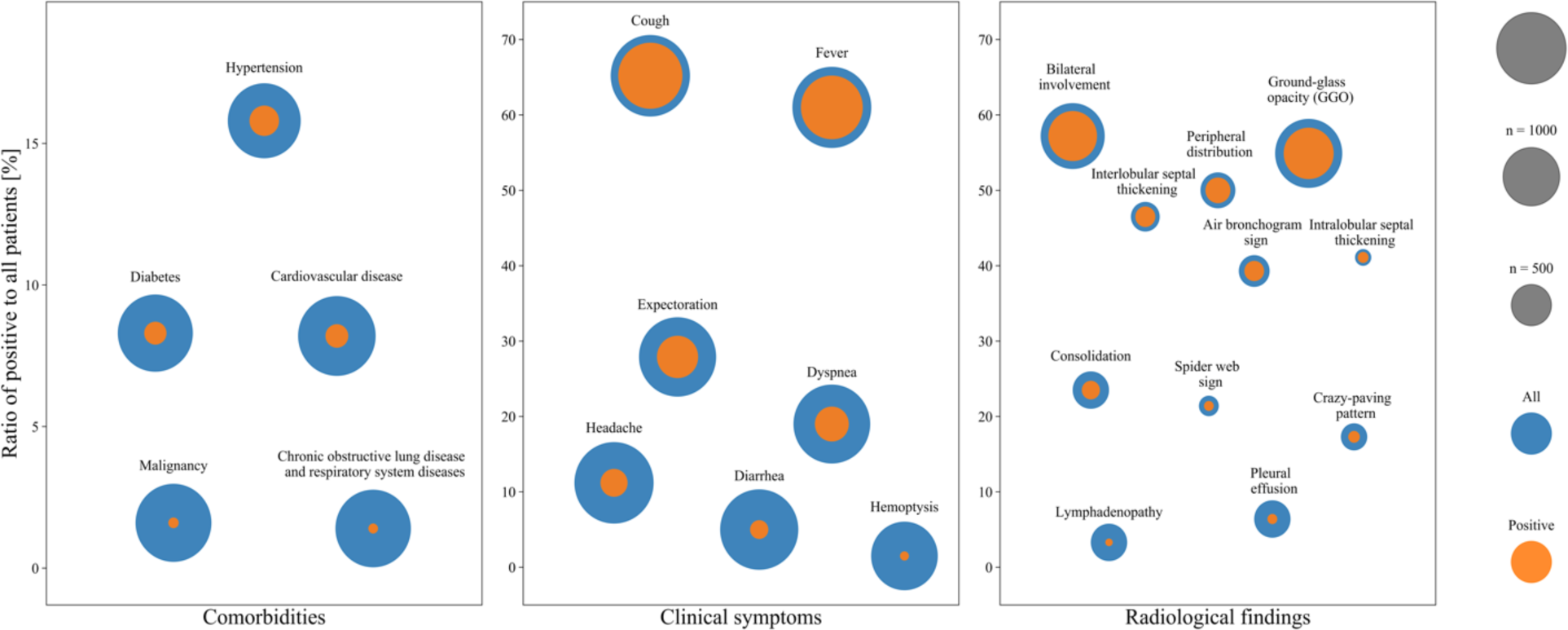
Evidence-based template generation. Pooled patient data from the reviewed literature regarding comorbidities, clinical symptoms and radiological findings of COVID-19 positive patients. The blue circles represent the total number of patients of which the parameter was analyzed. The inner orange circle represents the number of patients that fulfilled the respective parameter.

### An approach for unified data collection and analysis using comparable criteria

Our proposal emphasizes the importance of quality, integrity and traceability of data to allow for later epidemiological analysis and application of artificial intelligence algorithms. We wish to establish an optimized digital signature of the disease consisting of imaging biomarkers, patient history and other clinical follow-up data obtained in a reproducible matter. We demonstrate, that such a standardized diagnostic procedure can be easily and globally implemented. The proposed template or eCRF, respectively does not only provide a structured method for evaluating the pulmonary involvement but is also essential for quantitative assessment of the progression of the disease in clinical trials beginning in the near future. A standardized assessment is only possible if the imaging findings are quantitatively collected and evaluated on a supra-regional, preferably global level using comparable criteria. A representative assessment of COVID-19 pneumonia will be enabled by using the collected annotated data consisting of primary imaging data and secondary clinical data. These provide structured primary data that can be universally shared and analyzed. Therefore, an electronic case report form was developed on the basis of the mint lesion^™^ medical software product platform (Mint Medical GmbH, Heidelberg, Germany) (*Figure 2, 3*).

**Figure 2:**
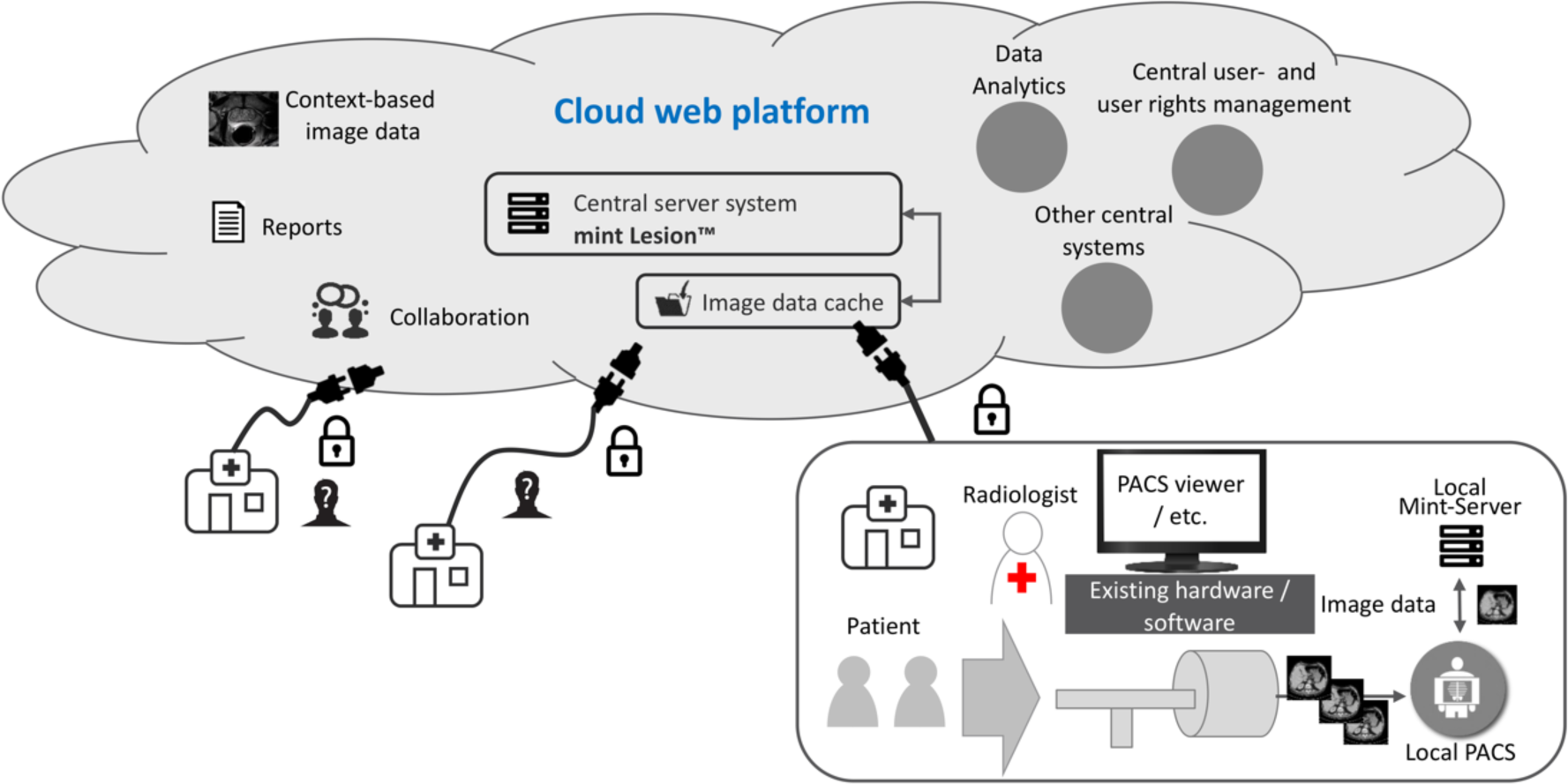
System. architecture cloud-based web platform. Every health care center has the possibility upload anonymized data to the web platform and use the eCRF based on the mint lesion medical product software package.

**Figure 3:**
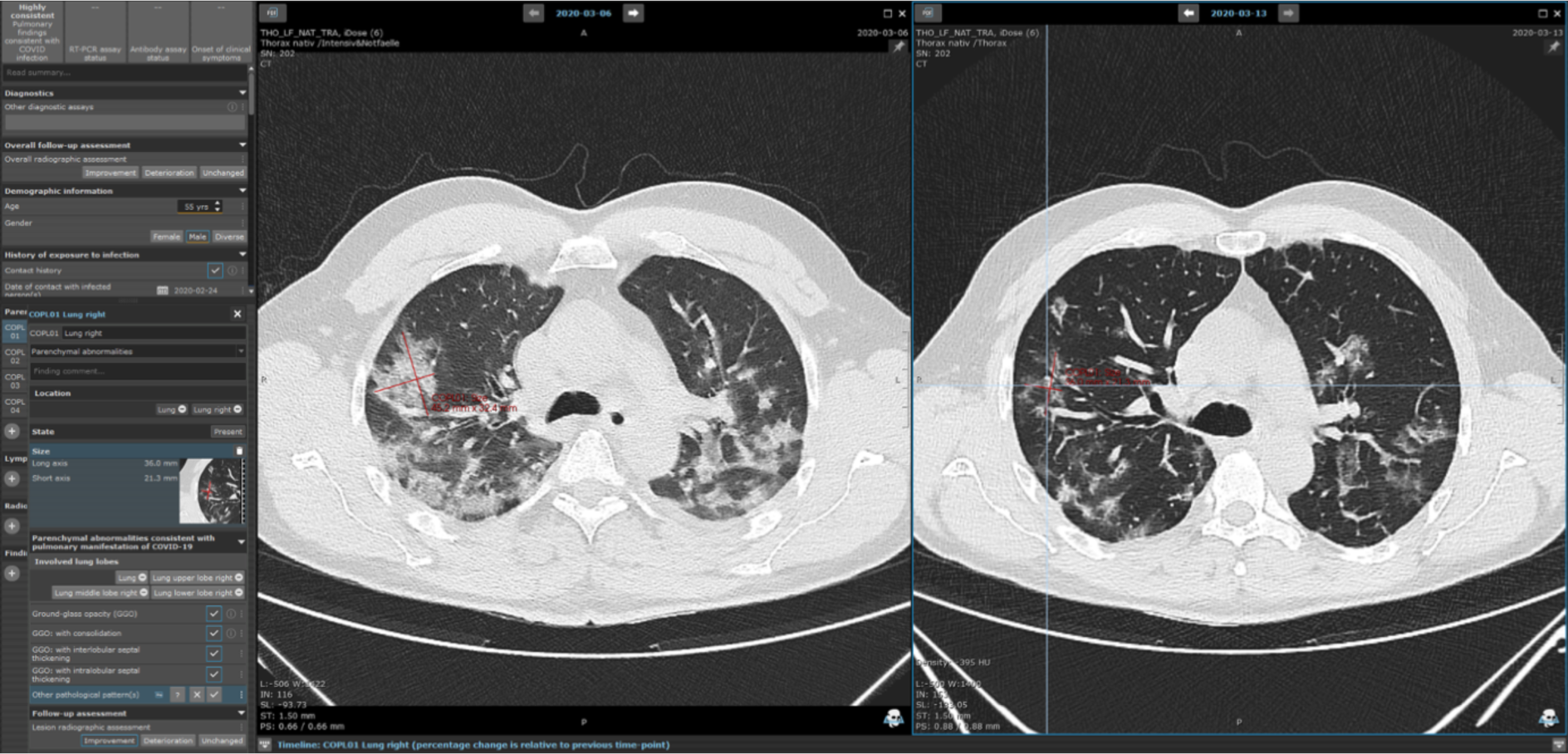
COVID-19 electronic case report form on mint lesion^™^ software platform. *Here:* Exemplary patient with confirmed COVID-19 pneumonia and chest CT at two subsequent time points. The patient was assessed and evaluated using the eCRF by an experienced radiologist. The assessment and evaluation are performed directly on primary imaging data and automatic rule-based evaluation of the disease progression. The patient was included in a clinical trial directly without the necessity of further data collection or twofold data documentation.

The template is available free of charge and has already been deployed to university hospitals and health care providers in 8 countries around the world, including severely affected regions of the United

States and Italy as well as the majority of the university hospitals in Germany. In addition, a cloud-based web platform will be provided, where every other hospital and health care center may have free access and can upload, assess, report and document their cases (Figure 2, full exemplary report s. supplementary information). Every center may choose and is encouraged to upload their pseudonymized data in order to contribute to an open-access repository. An online dashboard of the software will perform an explorative data analysis in real-time to monitor and alert if applicable. In addition, a customized data analysis dashboard can be created by every user. All pseudonymized data records submitted to the cloud are open-access and may be used to enhance the knowledge of COVID-19 pneumonia and contribute to improved clinical care of COVID-19 patients in the near future.

### Electronic case report form (eCRF)

The eCRF enables data acquisition in different levels. In detail, on an eCRF document level information about the patient such as demographic information and exposure history but also about already performed diagnostic measures and their respective results are requested to be recorded by the user. In addition, secondary clinical data such as possibly relevant comorbidities, clinical symptoms and clinical chemistry are queried. The selection of parameters is based on current study evidence as described above. A dynamic flow of refinements and amendments is possible without the risk of diminishing the quality of previously collected data. Furthermore, most recent results in evidence-based markers can dynamically be integrated in the template. The assessment of the imaging part includes general pulmonary radiological findings such as pleural effusion or lymphadenopathy. Pulmonary findings that have been found to be compatible or in combination with other findings even indicative for COVID-19 are automatically documented in a separate category depending on the following described image findings. The eCRF allows for the documentation and measurement of specific findings or general longitudinal monitoring on primary data. Methodological consistency and conformity of the measurements are facilitated by the software architecture. Additional automatically computed radiomics features will provide secondary data derived from the image. The measurement of parenchymal abnormalities and the documentation of the grade of the lesion (e.g. ground-glass opacification) at each time point (in case of several examinations) is annotated to the lobular sublocation in the lung. In accordance with the evidence described above, peripheral distribution pattern of the lesions, bilateral lung involvement and other qualitative specifications of the lesions with regards to their sublocation are evaluated automatically with a rule-based template framework. This leads to an intelligent and simplified workflow supported by the logical constraints. A report for each time point and for the longitudinal observation of the patient is generated automatically and might be used for internal communication between departments (exemplary excerpt Figure 4; for full report s. supplementary information).

**Figure 4:**
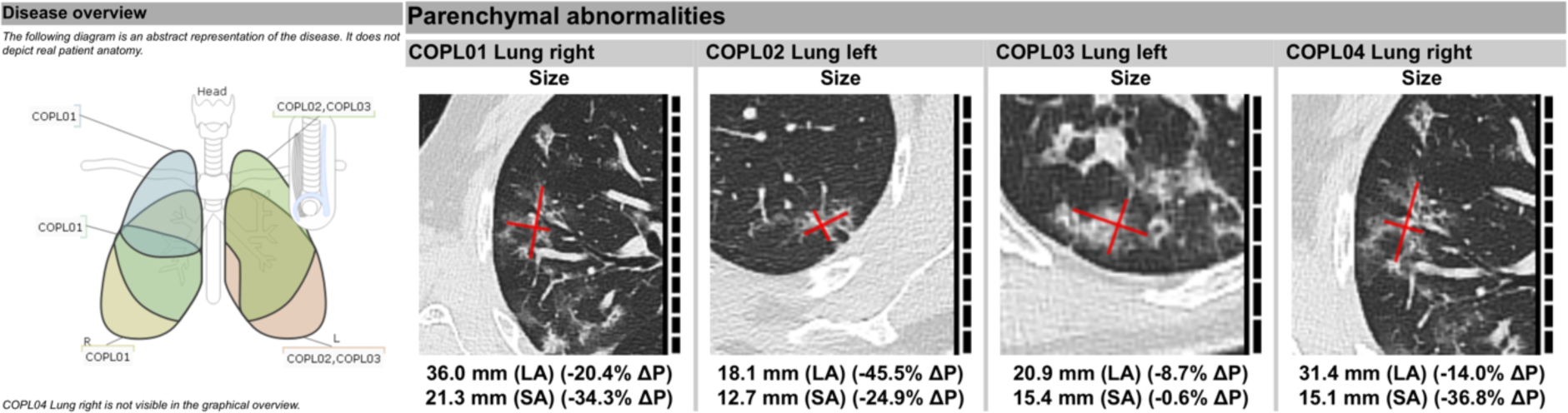
Overview of locations of major findings in chest CT. Longitudinal overview of documented findings in chest CT examination (*here:* evaluation after second chest CT in COVID-19 disease course). Recession of the size (LA: long axis, SA: short axis) of the imaging findings. These findings are included in an automatically generated case report file for COVID-19 positive patient and study data repository.

The particular strength of the developed eCRF lies in the fact that any imaging-based value is immediately linked to its context, e.g. anatomical location, but also further related data of clinical significance. Due to the integration in the mint lesion software platform minable data is obtained in a semantic data model with a HIPAA-and FDA CFR Part 11-ready audit trail, immediately applicable for Phase I-III clinical trials and other medical research. Furthermore, data is exportable in various human- and machine-readable data formats for external analysis. All image annotations can be exchanged in open formats, like NRRD, or as DICOM-compliant annotations (DICOM RT, DICOM Seg. Surface Obj.). Interfacing with other information systems, such as hospital information systems to include clinical data, is possible through standardized interfaces, including HL7, FHIR, and openEHR. These interfaces can also be used to transfer the automated report to the locally used RIS/HIS or other destinations.

## Discussion and Conclusion

The proposed solution supports the detection and standardized recording of clinical and radiological disease features consistent with the course of COVID-19 infection as well as the relevant epidemiological disease history. We envisage that this data-driven approach will be used in current and future research into therapies and vaccine candidates for COVID-19 pneumonia as well as future viruses pneumonias, fibrotic diffuse interstitial lung diseases or other diseases with complex characteristic patterns, as the evaluation of clinical trials on novel therapies and development of risk stratification markers can be performed using the provided eCRF template. In addition, future analysis can be performed on current and ongoing data reported via this structured approach due to the systematic and harmonized data collection. As of today, many of the published studies lack a sufficient number of cases or presented only retrospective results with partially incomplete information due to challenges in data acquisition. Thus, many studies conclude that there is a need for a broader and larger study with systematic data acquisition in the immediate future^10,12^. However, these studies provide first data eligible for inclusion in a systematic prospective study protocol. We created the Ecrf based on the data extracted from the current studies. As Radiologists become more familiar with radiological findings in line with COVID-19 pneumonia, the proposed eCRF will help to contribute substantially to diagnosis and treatment of COVID-19 infection and facilitate pooling of international knowledge. The template together with its automatically generated reports and computer-assistance might also serve as a learning platform for further education. The rapid, pandemic spread showed, that an extensive preparation and adjustment of healthcare systems is crucial to be able to respond appropriately and timely. We hope that the structured guidance through the radiological assessment together with the pooled information gathered from other sites might contribute to flatten the learning curve and thus improve clinical management of patients suffering from COVID-19 infection. Furthermore, resource scarcity in relation to the number of cases has been widely experienced by means of medical consumables and infrastructure but also a shortage of health care professionals. A cloud-based web platform dislodges the need for physical proximity to the hospitals or restrictions of quarantine measures of radiologists. Teleradiology services can be easily enabled based on the proposed concept, thus leading to larger capacities for COVID-19 pneumonia and beyond. Our approach has limitations. Data acquisition and handling of an eCRF is more time consuming than a reading in clinical routine. Further, integral data acquisition of imaging and clinical parameters requires an interdisciplinary input, which might not be available for every case. Currently, the template is designed for CT imaging only, but providing an eCRF for Chest-X-rays can be realized easily and is the scope for future work. Lastly, it might also appear controversial, that an increased research effort is encouraged even though manpower is limited during the current pandemic. The primary goal is to provide a tool for future clinical studies which are urgently needed.

We do not want to present a predefined global standard for the assessment and reporting for patients with COVID-19 infection, instead the intention is to initiate an evidence-based approach to define a joint global standard that can be adapted to the dynamic situations and discoveries yet to come. This might also discover regional difference in the disease leading to a personalized patient management. The eCRF can be adapted and refined according to evidence generated in ongoing and future clinical trials but also through the crowd-intelligence approach itself. Any addition or changes in template parameters will not put previously collected data in danger. Data integrity is permanently ensured, and even retrospective acquisition of certain parameters will be possible if applicable. Chest CT as a supporting tool for diagnosis and furthermore therapy monitoring has distinct benefits such as short turnaround time of the CT, wide availability of the equipment and the possibility of assessing the stage of the disease including response to therapeutic measures. However, it needs to be emphasized that even though the reporting is conducted directly on the CT image data, the eCRF can be used even if there is no imaging dataset available. Parameters regarding the patient history, clinical symptoms and clinical chemistry might still be documented systematically and shared with other ongoing activities like the leoss-project (https://leoss.net). Consequentially, the target group of users are not only radiological departments but every other department, that contributes to the diagnosis and treatment of COVID-19 patients. We aim to provide ‘role-dependent’ eCRF template solutions that can be integrated into one structured data set in the near future. Access to the cloud-based web platform and the open access data repository will be announced on https://mint-medical.com and through channels of the respective professional associations. With the presented systematic, computer-assisted and context-guided approach to capture data and annotations based on an easy-to-use software free of charge, we wish to contribute to a joint global response against the COVID-19 pandemic in a balancing act between research and clinical routine and further to initiate a change of clinical study culture.

## Data Availability

The manuscript itself is a report about a developed tool for collaborational data acquisition using standardized criteria for assessment, reporting and analysis of patients with COVID-19 pneumonia. It does not contain any original data, besides those included in the manuscript.

## Acknowledgements

We wish to thank the whole team of Mint Medical GmbH Heidelberg including H.G. Kenngott, M.D., E. Atsiatorme, M.D., A. Steinmann, M.D., J. Kast, Ph.D. and K.H Luby for their past and ongoing effort to supply every health care specialist with the developed software tool.

## References

1. Li R, Pei S, Chen B, et al. Substantial undocumented infection facilitates the rapid dissemination of novel coronavirus (SARS-CoV2). Science 2020.

2. Hu Z, Song C, Xu C, et al. Clinical characteristics of 24 asymptomatic infections with COVID-19 screened among close contacts in Nanjing, China. Sci China Life Sci 2020.

3. Hellewell J, Abbott S, Gimma A, et al. Feasibility of controlling COVID-19 outbreaks by isolation of cases and contacts. Lancet Glob Health 2020.

4. Lai CC, Liu YH, Wang CY, et al. Asymptomatic carrier state, acute respiratory disease, and pneumonia due to severe acute respiratory syndrome coronavirus 2 (SARS-CoV-2): Facts and myths. J Microbiol Immunol Infect 2020.

5. Bedford J, Enria D, Giesecke J, et al. COVID-19: towards controlling of a pandemic. Lancet 2020.

6. Fang Y, Zhang H, Xie J, et al. Sensitivity of Chest CT for COVID-19: Comparison to RT-PCR. 268 Radiology 2020:200432.

7. Guanmin Jiang XR, Yan Liu, Hongtao Chen, Wei Liu, Zhaowang Guo, Yaqin Zhang, Chaoqun Chen, Jianhui Zhou, Qiang Xiao, Hong Shan. Application and optimization of RT-PCR in diagnosis of SARS-CoV-2 infection. medRxiv 2020.

8. Ai T, Yang Z, Hou H, et al. Correlation of Chest CT and RT-PCR Testing in Coronavirus Disease 2019 (COVID-19) in China: A Report of 1014 Cases. Radiology 2020:200642.

9. Bernheim A, Mei X, Huang M, et al. Chest CT Findings in Coronavirus Disease-19 (COVID-19): Relationship to Duration of Infection. Radiology 2020:200463.

10. Guan WJ, Ni ZY, Hu Y, et al. Clinical Characteristics of Coronavirus Disease 2019 in China. N Engl 277 J Med 2020.

11. Liu K, Fang YY, Deng Y, et al. Clinical characteristics of novel coronavirus cases in tertiary hospitals in Hubei Province. Chin Med J (Engl) 2020.

12. Huang C, Wang Y, Li X, et al. Clinical features of patients infected with 2019 novel coronavirus 281 in Wuhan, China. Lancet 2020;395:497–506.

13. Wang D, Hu B, Hu C, et al. Clinical Characteristics of 138 Hospitalized Patients With 2019 Novel Coronavirus-Infected Pneumonia in Wuhan, China. JAMA 2020.

14. Wei J, Xu H, Xiong J, et al. 2019 Novel Coronavirus (COVID-19) Pneumonia: Serial Computed Tomography Findings. Korean J Radiol 2020.

15. Dai WC, Zhang HW, Yu J, et al. CT Imaging and Differential Diagnosis of COVID-19. Can Assoc 287 Radiol J 2020:846537120913033.

16. Wu J, Wu X, Zeng W, et al. Chest CT Findings in Patients with Corona Virus Disease 2019 and its Relationship with Clinical Features. Invest Radiol 2020.

17. Xu X, Yu C, Qu J, et al. Imaging and clinical features of patients with 2019 novel coronavirus SARS-CoV-2. Eur J Nucl Med Mol Imaging 2020.

18. Zhu W, Xie K, Lu H, Xu L, Zhou S, Fang S. Initial clinical features of suspected Coronavirus Disease 2019 in two emergency departments outside of Hubei, China. J Med Virol 2020.

19. Yang W, Cao Q, Qin L, et al. Clinical characteristics and imaging manifestations of the 2019 novel coronavirus disease (COVID-19):A multi-center study in Wenzhou city, Zhejiang, China. J Infect 296 2020.

20. Chen N, Zhou M, Dong X, et al. Epidemiological and clinical characteristics of 99 cases of 2019 novel coronavirus pneumonia in Wuhan, China: a descriptive study. Lancet 2020;395:507–13.

21. Xu YH, Dong JH, An WM, et al. Clinical and computed tomographic imaging features of novel coronavirus pneumonia caused by SARS-CoV-2. J Infect 2020.

